# One Health Index Calculator for India: Using Empirical Methods for Policy Stewardship

**DOI:** 10.1101/2024.07.25.24310983

**Authors:** Saveetha Meganathan, Arpit Katiyar, Esha Srivastava, Rakesh K Mishra

**Affiliations:** Tata Institute for Genetics and Society, Bengaluru, 560065, India; Centre for Cellular and Molecular Biology, Hyderabad, 500007, India; Academy of Scientific and Innovative Research (AcSIR), Ghaziabad 201002, India

**Keywords:** One Health, One Health Index Calculator, Fuzzy Extent Analysis, Modified Entropy-based Weightage Method, Policy Stewardship

## Abstract

**Background:** *One Health* is a collaborative approach that can be used to evaluate and enhance the sectors of human health, animal health and environmental health and emphasize their sectoral interconnectedness. Empirical evaluation of the one health performance of a country in the form of an index provides direction for actionable interventions such as targeted funding; prioritized resource allocation; rigorous data management and evidence based-policy decisions, amid other efforts such as public engagement and awareness on disease management; environmental degradation and preparedness towards disease outbreaks and thereby strengthening global health security. Thus, developing a One Health Index (OHI) Calculator for India is a significant step towards evidence based one health governance in the context of low-and middle-income countries.

**Objectives:** a) To develop a One Health Index Calculator for India with efficient and ease of use weighting methods and demonstrate the calculation of the One Health Index. b) To develop India-country-specific datasets through secondary data collection from reliable data sources. c) To determine the data gaps for policy stewardship.

**Methods:** We propose a One Health Index calculator to measure the One Health Index from an Indian context by adopting the Global One Health Index framework that comprises of 3 categories, 13 key indicators, 57 indicators and 216 sub-indicators. A secondary data collection was conducted to create a dataset for India from reliable sources. For measuring OHI, we have demonstrated two mathematical weightage methods, an efficient expert-based rating using Fuzzy Extent Analysis (FEA) and Modified Entropy-based Method (MEWM).

**Results:** We demonstrate the step-by-step OHI calculation by determining indicator scores using both FEA and MEWM weightage methods. Through secondary data collection an India-country-specific dataset has been created from reliable sources. From the datasets for India, the indicator values for 156 out of 216 sub-indicators were found available, while there was lack of data for the remaining 60 indicators. Further, a pilot correlation analysis was performed between 20 indicator scores and the relevant budget allocations for the years 2022-2023, 2023-2024, and 2024-2025. It was found that the increase in the budget over consecutive years has shown an increase in indicator score or better performance and vice versa.

**Conclusions:** The demonstrated OHI calculator has the scope to serve as a governance tool, while promoting data transparency and ethical data management. There is a need for a collaborative data federation approach that can resolve data gaps in the form of incomplete, missing and unavailable data. Further the scope of performing correlation analysis between budgetary allocation and performance of indicators gives empirical evidence for policymakers to improve intersectoral communication, multi-stakeholder engagement, concerted interventions, and informed policy decisions for resource allocation.

## Introduction

One Health is a participatory, collaborative approach to enhance the health of people, animals, and ecosystems over time. It recognizes the interdependence of the health of humans, domestic and wild animals, plants, and the larger environment. While health, food sources, water, energy, and the environment are all broad topics with sector-specific concerns, cross-sectoral and cross-disciplinary collaboration helps to protect human health, address health challenges such as the emergence of infectious diseases, antimicrobial resistance, food safety, and promote health and integrity of our ecosystems. The One Health approach has potential to address the complete spectrum of disease control, from prevention to detection, readiness, response and management while also contributing to global health security by making sense of the multisectoral interconnectedness and their impact on one another [1].

The term ‘One Health’ evolved during severe acute respiratory syndrome (SARS) 2003-2004 and H5N1 influenza (bird flu) outbreaks, highlighting the interconnectedness of human, animal, and environmental health. However, the ‘Manhattan Principles’ underscored this link, recognizing the need for collaborative approaches in global disease prevention [2]. These outbreaks emphasized the potential for unknown pathogens from wildlife, which led to the development of effective alert and response systems. Global cooperation, involving United Nations (UN), World Health Organization (WHO), Food and Agriculture Organization (FAO), World Organization for Animal Health (WOAH), United Nations Children’s Fund (UNICEF), and the World Bank, addressed the H5N1 outbreak with International Ministerial Conference on Avian and Pandemic Influenza (IMCAPI) playing a key role [3]. The primary drivers of the emergence of novel zoonotic infectious diseases include human activities, changes to ecosystems, land use, agriculture intensification, urbanization, international travel, and trade over the past three decades. These diseases, predominantly originating in wildlife, pose significant public health risks. The One Health approach is pivotal for preventing, monitoring, and surveilling zoonotic diseases, emerging infectious diseases, sustaining food security, and combatting antimicrobial resistance (AMR), all of which affect human, animal, and environmental health. Subsequently, collaborative monitoring systems are now recognized as essential for effectively managing pandemics and outbreaks, given the multitude of epidemics, pandemics, and outbreaks in the last decade [3].

Developing a One Health index framework can contribute to improving the implementation of the One Health approach by focusing on intersectoral collaborations and their corresponding datasets. The value of such an index extends beyond compiling data; it has the potential to revolutionize our understanding, management, and actions concerning the complex web of factors that influence global health. A One Health index simplifies health data, making it accessible to diverse stakeholders including government functionaries, policymakers, healthcare providers and the public [4]. It condenses a huge volume of information into a quantifiable value. Government and healthcare organizations can utilize health indices to make informed policy decisions regarding the allocation of resources and implementation of public health interventions [5]. Further such indices facilitate the monitoring of health trends of a given region or country, enabling the assessment of the effectiveness of public health campaigns; improvements in healthcare systems and policy frameworks [6], [7].

In this milieu, the Global One Health index (GOHI) serves as an empirical tool for the systematic assessment of the One Health scores of more than 200 countries/territories globally [8]. The GOHI framework consists of three major categories: extrinsic driver index, core driver index and intrinsic driver index which are further categorized into 13 key-indicators, 57 indicators and 216 sub-indicators, thus making it possible to quantify a One Health index for a country by mapping multisectoral variables contributing to the well-being of humans, animals and environment and their impact on one another [8]. Additionally, the GOHI framework accounts for the recruitment of 29 domain experts to attribute weightage to the indicators based on their sectoral experience, further demonstrates the need for cross-sectoral communication and concerted efforts to achieve better one health outcomes for a country [8].

In this study, we have espoused to adapt and contextualize the One health Index calculator for India from the GOHI framework. We have demonstrated the calculation of the indicator scores using two mathematical weightage methods: i. the efficient expert method which is the Fuzzy Extent Analysis (FEA) that requires recruitment of domain experts and ii. the Modified Entropy-based Weightage method (MEWM), which replaces expert-based weightage calculation with mathematical formulae-based calculations. A country specific database for India has been developed through secondary data collection. The two mathematical weightage methods – FEA and MEWM are demonstrated using the India specific database to generate indicator scores, subject to data availability. In addition, we have corelated the sectoral budget over the last two financial years with the sectoral indicator scores and this serves as an actionable pointer for policy makers, federal and state governments in decision making towards budgetary allocations.

## Methodology

The One Health Index calculator for India is aimed to serve as a public health tool and address the need for multisectoral collaboration for data access and to understand the impact of sectoral performance and thereby improve the countrywide ‘One Health’ performance. This calculator consists of the following steps:

### Indicator selection

For demonstrating the calculation of One Health Index (OHI) for India, we are adopting the list of 3 categories, 13 key-indicators, 57 indicators and 216 sub-indicators and the weightages assigned to each of them from GOHI [8] (Multimedia Appendix 1).

### Database building

For the demonstration of the One Health Index Calculator for India, the data collection has been done using secondary data sources such as Press Information Bureau (Government of India), Department of Agriculture and Farmers Welfare (Government of India), World Bank, Food and Agriculture Organization of the United Nations, Yale Environmental Performance Index (Yale University), Our World in Data, The Global Economy, Statista, India Stat, Knoema and other relevant databases (Multimedia Appendix 2). Further, to corelate the sectoral budget over the last two financial years with their respective indicator scores, the budget datasets were obtained from the Indian Union budget document for the financial years 2022-2023, 2023-2024, and 2024-2025 (Multimedia Appendix 3).

### Weight determination

A sample proforma has been developed to obtain expert ratings for different sub-indicators, indicators, key-indicators, and categories. This proforma can be self-administered if shared via e-mail or can be used for in-person interviews and thereby provides ease of use for data collection. The proforma allows the expert to provide additional information like variables, data sources and case studies. (Multimedia Appendix 4).

### Procedure to calculate expert weights

There are two mathematical weightage methods for calculating the indicator scores:

a. the efficient expert-based method which is the Fuzzy Extent Analysis (FEA)
b. the Modified Entropy-based Weightage Method (MEWM)

#### Fuzzy Extent Analysis (FEA)

The efficient expert-based rating method to calculate the indicator scores will require consultations with experts from diverse One Health sectors for ascertaining the priority or the weightage of different sub-indicators and indicators. The Fuzzy Extent Analysis (FEA) is a multi-criteria decision-making method that integrates both qualitative and quantitative approaches [9]. Data collection for expert-based rating can be done using the given proforma. For developing the proforma, a pair-wise comparison was used for the experts to rate the metrics [10]. It is an effective way to gather opinions from many experienced experts, especially for complex decision-making problems involving multiple risks. The following is an example of how expert based ratings will be converted into weights. A pair of sub indicators belonging to the experts’ domain area will be provided to them for pairwise comparison through linguistic ratings, for which there already exists a numerical scale of relative importance [10]. Similarly, all possible combinations of sub indicators will be provided to the expert for comparisons. The linguistic ratings obtained from an expert, such as-equally important, moderately important, strongly important, and extremely important will be converted into numerical ratings. These numerical ratings will be further converted into comparison matrix. Further, the consistency of the responses by the experts will be checked using the consistency ratio. After ascertaining the consistency of the responses by the experts, the next step will be fuzzification (to convert the numerical ratings to fuzzy numbers) of the ratings. To convert the numerical ratings to fuzzy numbers, we will use triangular fuzzy numbers (a generalization of real numbers, representing a set of possible values with weights, or membership functions) to calculate the weightage of different sub-indicators and indicators. Similarly, the weights for the key indicators and categories can be collected using the efficient expert-based weightage mechanism or a format like a panel discussion can be conducted to obtain the weights from the domain experts.

#### Steps involved for OHI calculation using Fuzzy Extent Analysis (FEA)

*Step 1:*

a. Consult with domain experts from diverse sectors relevant to One Health.
b. Based on the systematic implementation of the semi-structured interviews and a modified Delphi method, pairwise comparisons are made on the importance of each pair of parameters.
c. Consider a hierarchy with n parameters, all of which must be n(n-1)/2 pair-wise comparisons.
d. The comparisons are rated linguistically.
e. Linguistic variables are converted to numerical ratings using scale of relative importance [11].

*Step 2:*

A comparison matrix will be established for each expert using the ratings from the proforma. The comparison matrix for the *k*^*th*^ expert is given by, 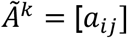, that is represented as:

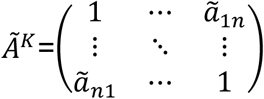

The reciprocal of the matrix is denoted by 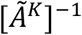.

The properties of reciprocity can be given as

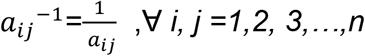

*Step 3:*

Consistency Index calculation

It is crucial to maintain consistency for the expert weightage process to obtain the indicator scores. To achieve this, a Consistency Index (CI) was introduced to guarantee the consistency of the comparison matrix [10].

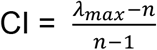

where, *λ*_*max*_ is the maximum eigen value and n being the dimension of the comparison matrix [9].

The consistency ratio is given by:

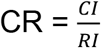

Where, RI being the random consistency index which depends on *n* [10].

|  | 1 | 2 | 3 | 4 | 5 | 6 | 7 | 8 | 9 | 10 |
| --- | --- | --- | --- | --- | --- | --- | --- | --- | --- | --- |
| RI | 0 | 0 | 0.52 | 0.89 | 1.12 | 1.26 | 1.36 | 1.41 | 1.46 | 1.49 |

If CR < 0.1, then the judgement is consistent [12].

*Step 4:*

a. Convert the ratings in the comparison matrix to triangular fuzzy number.
b. A fuzzy number serves as a tool to express values that are uncertain or imprecise particularly in the context of fuzzy set theory.
c. Unlike a value, it accommodates varying degrees of belongingness enabling it to account for the ambiguity often found in practical scenarios.
d. Fuzzy sets can compensate for the inconsistency and imprecisions in human judgements rather than random or stochastic ones.
e. Fuzzy numbers are depicted by a set of possible values having their own membership function ranging from 0 to 1. with the member function as:
f. A triangular fuzzy number is represented by [floor value, average value, ceiling value] i.e., (*l*, *m*, *u*) with the member function as:

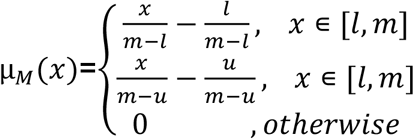

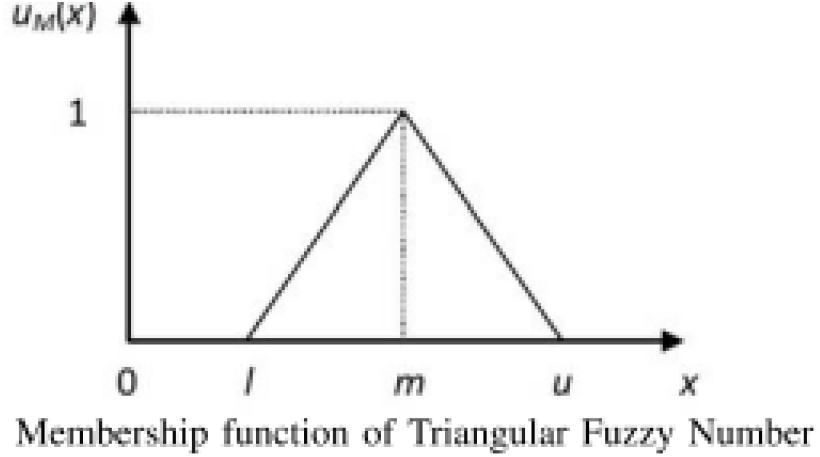

[13]

*Step 5:*

a. For calculating the weights from a comparison matrix, fuzzy extent analysis is pertinent [13].
b. In Extent Analysis X={*x*_1_,*x*_2_,…,*x*_*n*_} being an object set and G={*g*_1,_*g*_2,…,_*g*_*n*_} being a goal set. Then for each object, extent analysis is performed correspondingly for each goal *g*_*i*_.

The value of fuzzy synthetic extent is given by,

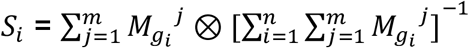

Where,

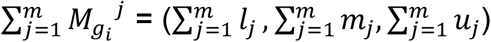

We can calculate the ratings in the triangular fuzzy number format as given in the above equations leading to a weight derived from these triangular weights.

A pairwise comparison of fuzzy weights needs to be performed and the computation of the degree of possibility of them being greater than the fuzzy weight will be obtained. The minimum of these possibilities is used as the overall score for each criterion *i*.

To compute the value of the fuzzy synthetic extent, *S*_*i*_ for the *i*^*th*^ object is as follows:

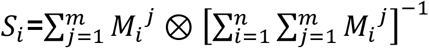

Where,

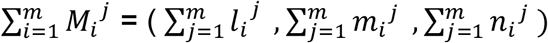

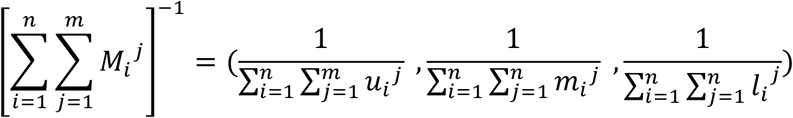

*Step 6:*

The fuzzy synthetic extent will constitute three values which will be averaged out to acquire a single value for the weights from them.

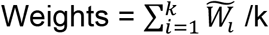

*Step 7:*

Normalize the weight by dividing the individual weights by the sum of all weights.

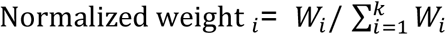

*Step 8:*

If the ratings have been collected from multiple experts for the same indicator, then there are two methods to get the optimized ratings:

*Method 1:*

Step a: Calculate individual normalized weights using all the above steps for every expert rating.

Step b: Average the normalized weights by the experts to acquire optimized ratings for the indicators.

*Method 2:*

In continuation to step 5, after calculating the fuzzy synthetic extent for every expert input individually,

i. Compute the degree of possibility of

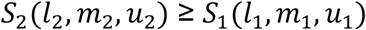 Where degree of possibility between two fuzzy synthetic extents is defined as

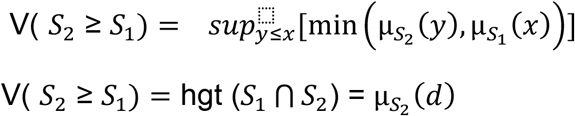 Where,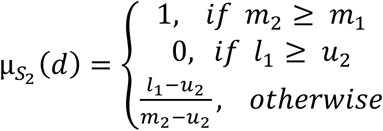 And d is the ordinate of the highest intersection point d between 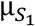 and 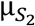.

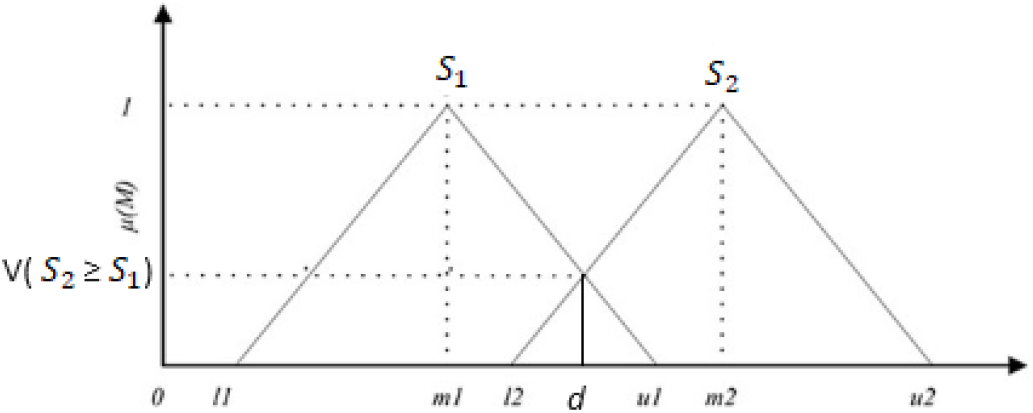
ii. Compute the degree of possibility for a convex fuzzy number to be greater than k convex fuzzy numbers *S*_*i*_ (1,2,…,k) V [(S ≥*s*_*1*_ , *s*_*2*_ ,…, *s*_*k*_ ) =V [(S ≥ *s*_*1*_ ) and (S ≥ *s*_*2*_ ) …and (S ≥ *s*_*k*_ )] = min V (S ≥ *S*_*i*_ ), i=1, 2, …, k
iii. Compute the vector W’.

W’ = (*d ′*(*A*_1_),*d ′*(*A*_2_),…, *d ′*(*A*_*k*_))^*T*^

Where d’(*A*_*i*_) = min *V*(*S*_*i*_ ≥ *S*_*j*_) for i =1, 2, …, k and j=1, 2, …, k and i ≠ j

Normalized vector, W = (*d*(*A*_1_),*d*(*A*_2_),…,*d*(*A*_*k*_))^*T*^

W is a non-fuzzy number calculated for each comparison matrix [14]

Now, the weights have been calculated through Fuzzy extent analysis (FEA), thus using these weights and an appropriate weight accumulation formula One Health Index can be calculated.

*Step 9:*

For the accumulation of the indicators and the weights, the following formula can be used:

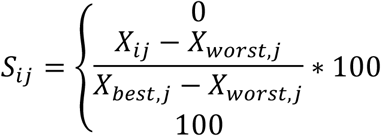

Where *S*_*ij*_ denotes the normalized score for the *j*^*th*^ sub indicator of *i*^*th*^ indicator; *X*_*ij*_ denotes the original values for the *j*^*th*^ sub indicator of *i*^*th*^ indicator, *X*_*best*,*j*_ denotes the best value for the *j*^*th*^ sub indicator of the *i*^*th*^ indicator, *X*_*worst*,*j*_ denotes the worst value for the *j*^*th*^ sub indicator of the *i*^*th*^ indicator. In cases where no data is available for the sub-indicators, substitute *S*_*ij*_ with 0.

The weighted sum of the scores can be given by:

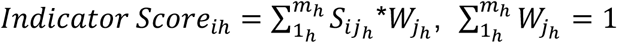

Where m depicts the number of sub indicators under the *h*^*th*^ indicator, *j*_*h*_ depicts the *j*^*th*^ sub-indicator under the *h*^*th*^ indicator;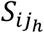 depicts the score of the *j*_*h*_ ^*th*^ sub indicator under *i*^*th*^ indicator;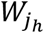 depicts the weight of *j*_*h*_ ^*th*^ sub indicator [8].

The procedure needs to be repeated stage-wise for sub-indicators, indicators, key-indicators and categories to finally calculate One Health Index.

#### Modified Entropy based weightage method (MEWM)

a. This approach requires the indicators values for OHI calculation.
b. Weightage mechanism is backed by the entropy-based weightage method for the indicator value and is a preferred method due to the ease of calculation and does not require expert-based rating.

*Step 1:*

First, normalization (a systematic process of organizing data in a database to make it more flexible and cohesive) of the values of the sub indicators will be done using the following formula,

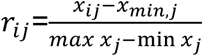

Where max *x*_*j*_ and min *x*_*j*_ are the maximum and minimum values among the alternatives for indicator j [15] .

*Step 2:*

The entropy *E*_*ij*_ of each sub-indicator i from the indicator j, the entropy *E*_*ij*_ of eachindicator was determined from the normalized values *r*_*ij*_ as formulated:

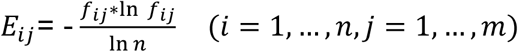

Where 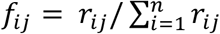.

For the cases where *f*_*ij*_ = 0 or not available, the entropy becomes not defined and, in those cases, substitute the entropy with 0.

*Step 3:*

For the applicability of the method, the weights *w*_*ij*_ are computed as defined below:

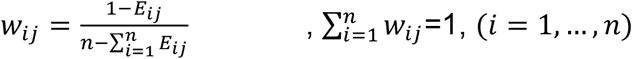

*Step 4:*

The above formula can be used at every step to calculate the relevant weights,

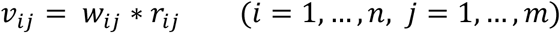

indicator value.

Where *r*_*ij*_ is the standardized value of the *i*^th^ sub-indicator for the *j*^*th*^ state and *w*_*ij*_ being the weight calculated for the *i*^*th*^ sub-indicator for the *j*^*th*^ state. *v*_*ij*_ is the weighted indicator value.

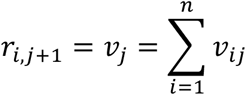

Where *v*_*ij*_ being the weighted indicator value for the *i*^*th*^ sub-indicator for the *j*^*th*^ state and *v*_*j*_ being the index value of the indicator j using which, calculate *r*_*j*_.

i. One can use this formula to arrive at the weighted value of the sub indicators and then use those sub-indicator values to derive weights for the indicators and similarly repeating the process for all available data.
ii. These steps will transition stagewise in an agglomerative way from the scores of the sub indicators of a particular indicator to the One Health index.

## Results

Using secondary data sources, a country specific dataset for India was developed. From this dataset, the indicator values for 156 sub-indicators out of 216 sub-indicators were gathered, while there was lack of data for the remaining 60 sub-indicators (Figure 1 and Multimedia Appendix 5). The developed dataset for India reflects data gaps, that is, inconsistency in data availability and areas where there is absence of data requires planned interventions by the governance systems. For some cases where the current data is available but due to absence of historical data the score could not be computed.

**Figure 1.**
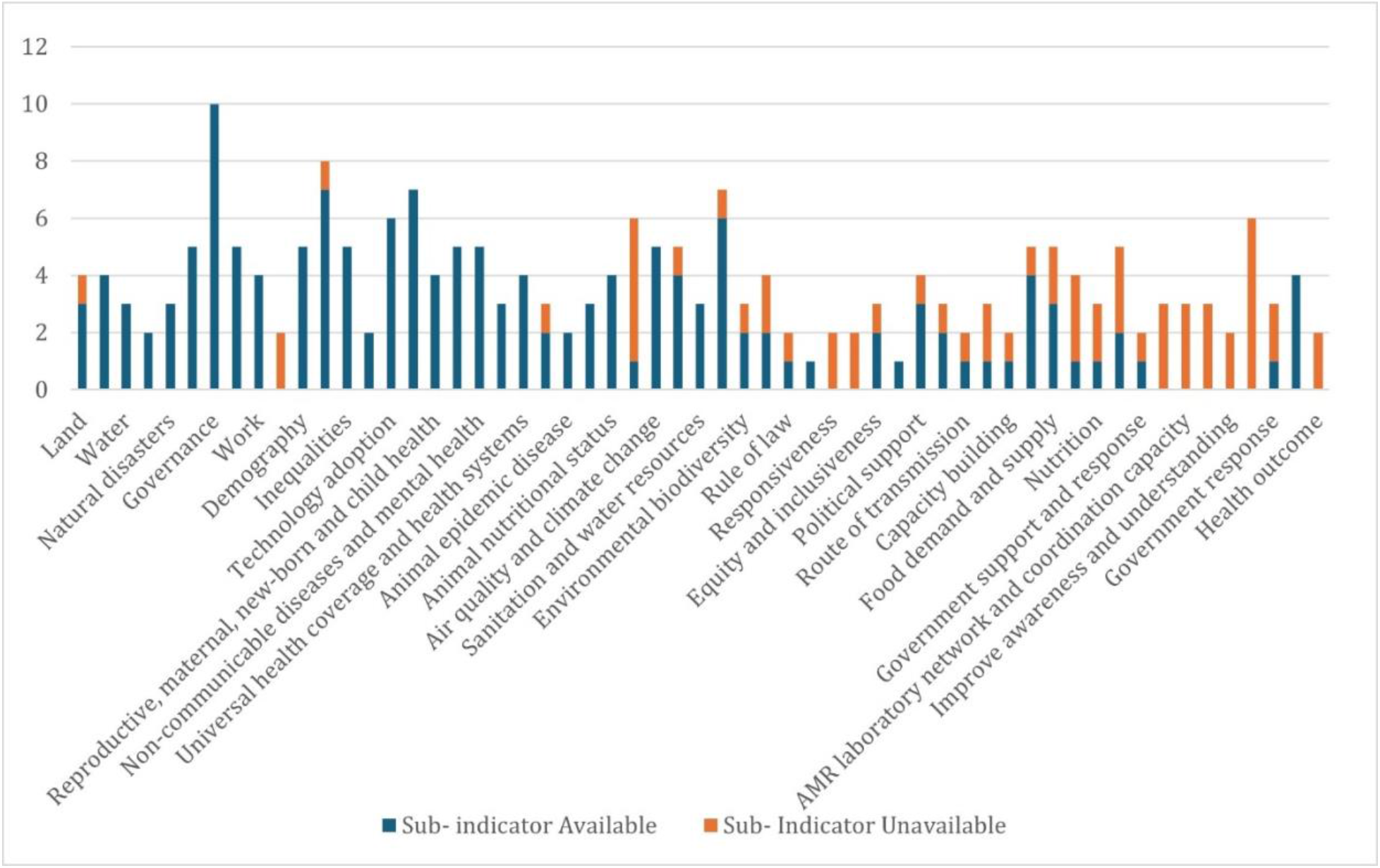
Data availability for the indicators. In blue is the number of sub-indicators for which the data is available and in orange is the number of sub-indicators for which the data is unavailable.

### Pilot Analysis

Additionally, to evaluate the efficiency of weightage methods, a comparative analysis was conducted between FEA and MEWM. Indicator scores were calculated for 23 indicators where data for all or most sub-indicators were available (Figure 2). Figure 2. shows that the indicator scores obtained using the two weightage methods, FEA and MEWM for 23 indicators are significantly consistent with one another and thereby rendering the two methods to be reliable. Further, to check the One Health calculator’s applicability and utility in the Indian context, a correlation analysis was performed between 20 indicator scores and budget allocations for the years 2022-2023, 2023-2024, and 2024-2025 (Multimedia Appendix 6). The differences in budget allocations for each consecutive year have been calculated, aiding in understanding the correlation between the indicator scores and the changes in budget allocations (Figure 3, 4). Table 1. shows that the increase in the budget over consecutive years has shown the rise in indicator score and vice versa.

**Table 1.** Correlation between the increase in budget and indicator scores.

| **Difference Between** | **FEA** | **MEWM** |
| --- | --- | --- |
| 2024-2025 and 2023-2024 | 0.36361013 | 0.23647898 |
| 2023-2024 and 2022-2023 | 0.311369994 | 0.20383558 |

**Figure 2.**
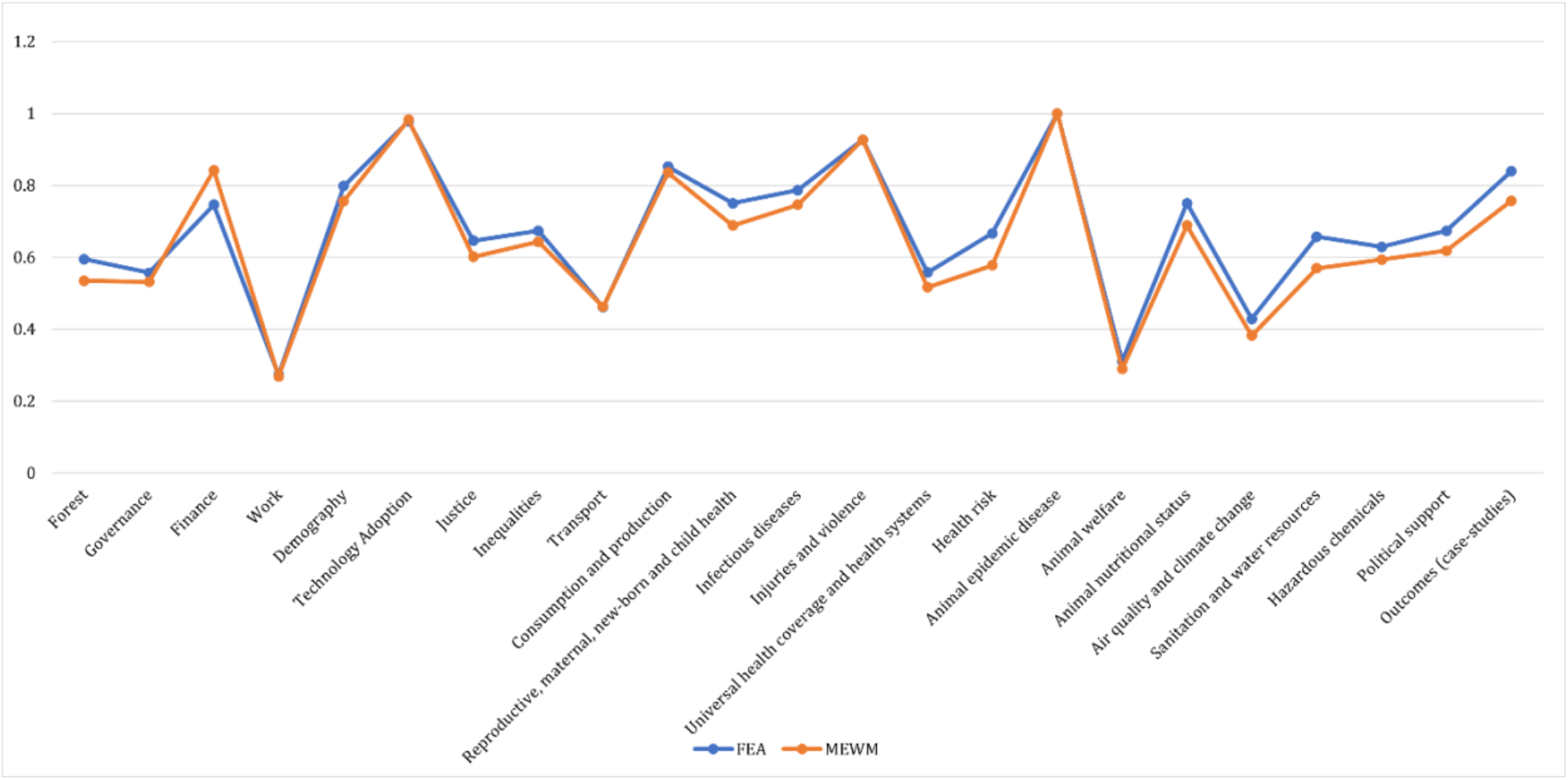
Indicator scores for different indicators using FEA vs MEWM

**Figure 3.**
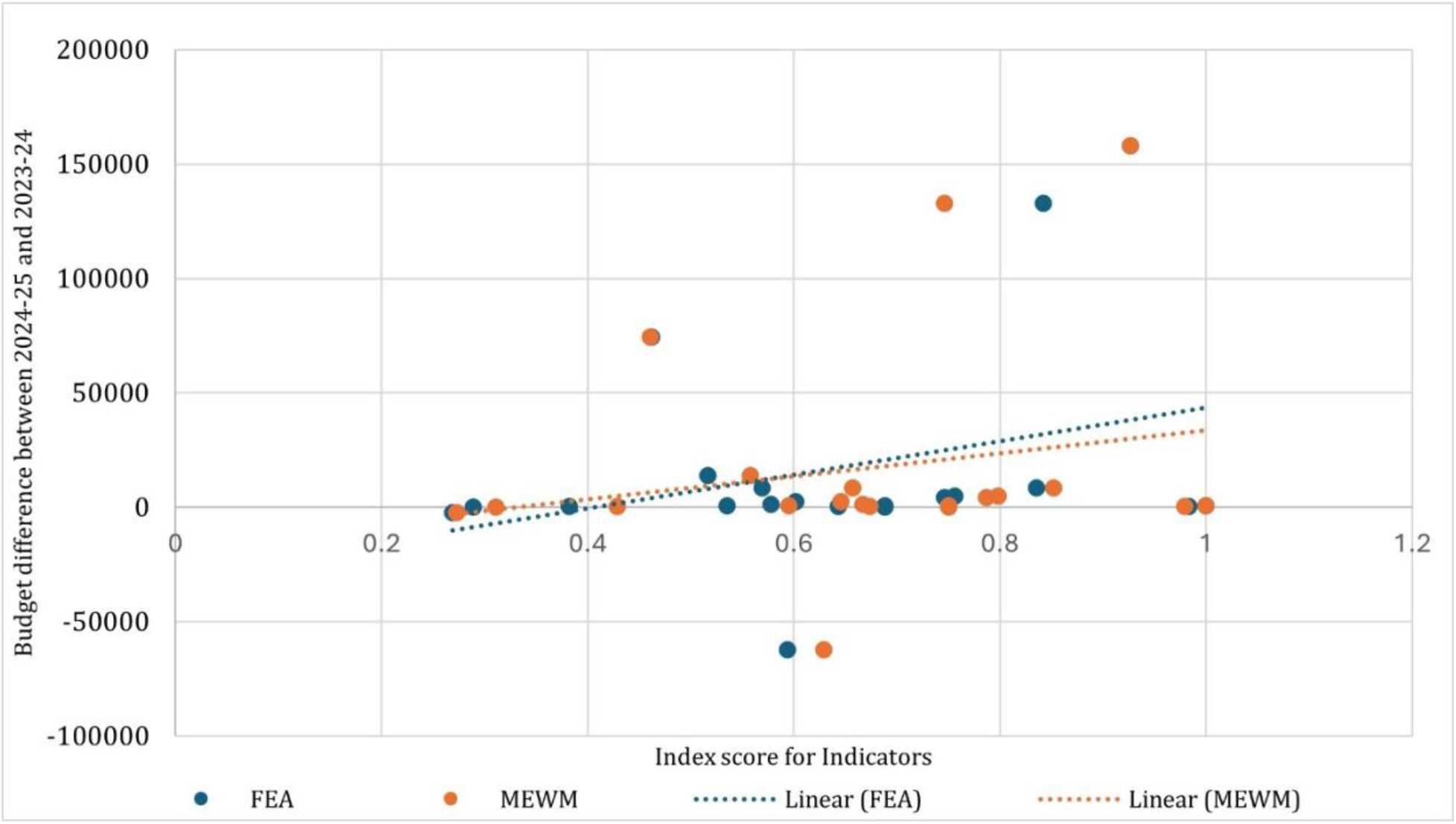
Budget difference of 2024-25 and 2023-24 against the indicator Index score. shows the budgetary differences for the years 2024-25 and 2023-24 against the indicator scores for consecutive years of budget allocations using the FAHP and MEWM methods.

**Figure 4.**
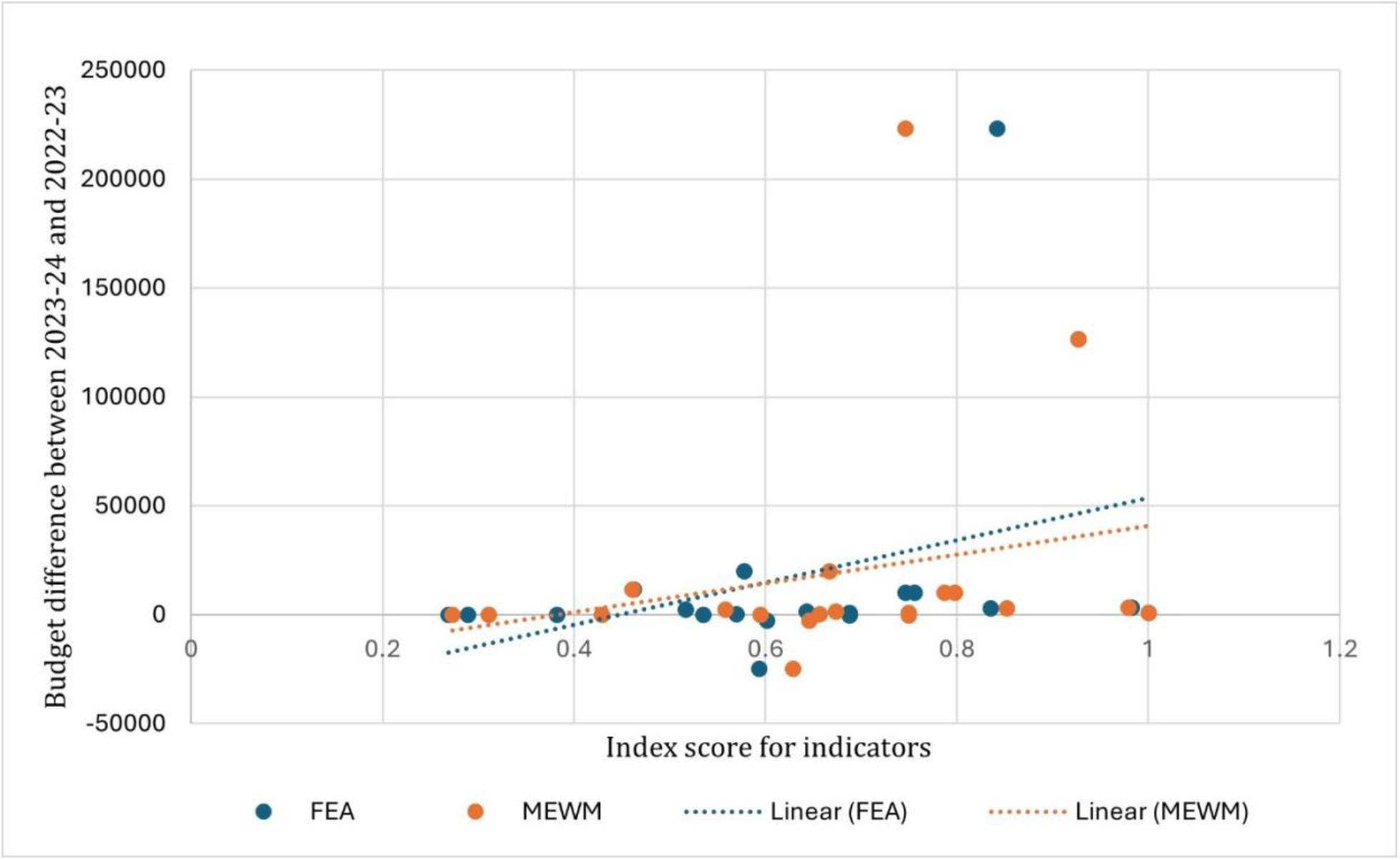
Budgetary differences for the years 2024-25 and 2023-24 against the indicator scores for consecutive years of budget allocations using the FAHP and MEWM methods. A positive correlation is observed for both methods, indicating that an increase in budget allocation positively impacts the indicator scores.

This emphasizes the importance of budgetary allocation in the performance of indicators thereby impacting the overall performance of One Health in India. The One Health index calculator has the potential for such nuanced correlation analysis to be performed, in this case, between budgetary allocation and the performance of the indicators through their scores. This can serve as a valuable insight for policymakers and stakeholders alike for prioritizing sectoral interventions related to One Health.

## Discussion

The One Health Index can be computed at various levels of governance in India which is constituted of 28 states and 8 Union Territories, depending upon data availability, such as national, state, district or even at block or village/panchayat levels. By calculating these values locally and then aggregating them, an accurate national value can be derived that highlights demographic variations and provides a more precise measurement. For state-level calculations, population density can serve as a key weighting factor. Collaborative monitoring systems are now recognized as essential for effectively managing pandemics and outbreaks [3]. Thus, developing a One Health index calculator can contribute to improving the implementation of policies using a participatory One Health approach. However, the value of such an index extends beyond compiling data; it has the potential to revolutionize our understanding, management, and actions concerning the complex web of factors that influence global health. The process of calculating a One Health Index for India sheds light on critical areas requiring systematic interventions, aided by policy decisions, particularly regarding resource allocation and strengthening of governance systems. This framework also promotes a collaborative data federation model to address data gaps— such as incomplete data, lack of timely data, and the absence of appropriate data—and correspondingly advocates for data transparency and ethical data management. Implementing a collaborative data federation model and maintaining consistent data collection will address these challenges and establish a historical dataset for indexing indicators. This approach encourages intersectoral communication, multi-stakeholder engagement, garners interest from governance systems, and builds momentum towards improving poorly performing indicators, thereby achieving better One Health outcomes for the country. A study focusing on mitigating zoonotic disease risk using the economic approach stated that allocating budget and resources strategically at a higher level secures sufficient funding to manage diseases along the livestock value chain, leading to improvements in human health [16]. As shown in Table 1, increasing the budget over consecutive years has led to higher indicator scores and vice versa, suggesting that budget increases for relevant ministries and departments contribute to improved indicator scores, thereby enhancing the OHI. Addressing the standardization of impact indicators and integrating new field knowledge are also essential.

In this study, we have provided a One Health Index Calculator tool, demonstrating the two weighting methods FEA and MEWM, alongside the India-specific datasets which is accessible and efficient of use for multiple stakeholders such as government functionaries; policy makers; researchers; and institutes of disease surveillance and preparedness, etc. While the scope of the study is to develop a One Health Index Calculator for India, and provide a reliable, ease of use and efficient tool that could be easily adapted by relevant sectoral stakeholders and compute empirical scores to plan informed and strategic interventions. We have also demonstrated the OHI value calculation for India using both the FEA method where we have assigned equal weightage in the place of expert weightage and the value obtained is 46.51 and the score obtained using the MEWM is 42.29 (Figure 5). These two demonstrated OHI calculation values are within the value range for South Asia region, which is 35-50 as published in 2022, using the GOHI framework [8]. Further, it is also pertinent to consider expert consultations to review the indicators as adopted from the GOHI framework and draw a list of indicators for India which may be region-specific and locally adapted.

**Figure 5.**
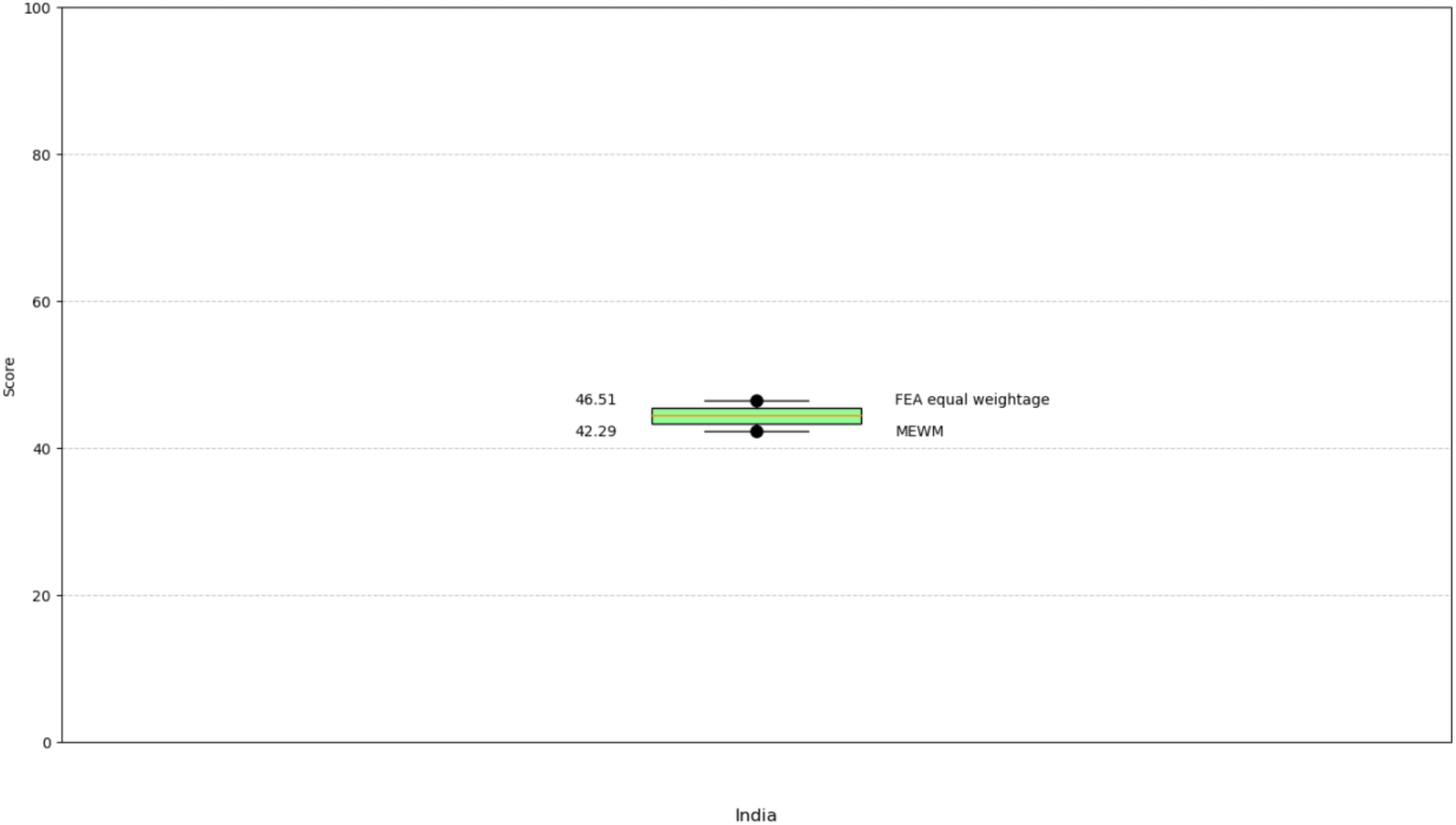
Demonstrated OHI calculation for India using both FEA (equal weightage) and MEWM

## Conclusion

The One Health Index (OHI) calculator identifies key areas that require concerted actions from One Health-centric stakeholders. Data gaps and deficiencies for crucial One Health indicators were identified during its development for data federation and open data access from governance systems and research organizations working on public interests emerges as an actionable agenda for concerted efforts. In this milieu, the methodologies and framework for calculating a single OHI value requires multi-sectoral experts to come together to work towards improving the One Health Index for India by recognizing the need for sectoral data. This in turn leads to identifying disparities, targeting interventions, monitoring health trends and other strategic efforts towards the health equity paradigm. Additionally, the importance of budgetary allocation for improving indicator scores that contribute to OHI is yet another reminder for policy makers towards empirical and evidence-based decision making. The process of calculating an empirical OHI value demands consistent public interface and awareness about the interconnected nature of the One Health approach and leads to better preparedness to handle future pandemics; improve quality of life and achieve sustainable development goals.

## Supporting information

Multimedia Appendix 1

Multimedia Appendix 2

Multimedia Appendix 3

Multimedia Appendix 4

Multimedia Appendix 5

Multimedia Appendix 6

## Data Availability

All data produced in the present work are contained in the manuscript

## Notes

### Competing Interest Statement

The authors have declared no competing interest.

### Funding Statement

The Tata Trusts

### Author Declarations

The source data are openly available and obtained using secondary data collection.

