## Supplementary material for "One Health Index Calculator for India: Using Empirical Methods for Policy Stewardship": Multimedia Appendix 4

**Sample Proforma for the Framework**

Name:

Area of Expertise:

Years of Experience (in the relevant field):

Considering the scale of relative importance of one sub-indicator in comparison to other sub-indicators we seek your preference.

Use the scale of relative importance for the comparisons given below:

| Scale of Relative Importance | Linguistic Rating |
| --- | --- |
| 1 | Equal importance |
| 2 | Intermediate between equal and moderate importance |
| 3 | Moderate importance |
| 4 | Intermediate between moderate and strong importance |
| 5 | Strong importance |
| 6 | Intermediate between strong and very strong importance |
| 7 | Very strong importance |
| 8 | Intermediate between very strong and extreme importance |
| 9 | Extreme importance |
| 2, 4, 6, 8 | Intermediate values |
| 1/2,1/3,1/4,1/5,1/6,1/7,1/8,1/9 | Values for inverse comparison |

If you rate your preference of one sub-indicator over the other sub-indicator, be any Natural Number other than 1 then the preference for the one which is less important against the important one will be inversed when compared against each other.

An **example** to demonstrate:

If Rohan likes an apple over a banana and he feel that an apple is 4 times important in comparison to banana for him then,

|  | Apple ✓ | ☐ | Banana | ☐ | 1 \| 2 \| 3 \| ✓4 \| 5 \| 6 \| 7 \| 8 \| 9 |
| --- | --- | --- | --- | --- | --- |

Then the rating of Banana in comparison to an apple for Rohan automatically converts to ¼.

For the Indicator Land, which do you see important over the other (mark the one with a ✓) and what level of importance does the preferred one hold over the other in One Health according to you?

| S.no. | Sub-Indicator-1 |  | Sub-Indicator-2 |  | Level of Importance |
| --- | --- | --- | --- | --- | --- |
| 1. | Country area | ☐ | Cultivated area | ☐ | 1 \| 2 \| 3 \| 4 \| 5 \| 6 \| 7 \| 8 \| 9 |
| 2. | Country area | ☐ | Arable land area | ☐ | 1 \| 2 \| 3 \| 4 \| 5 \| 6 \| 7 \| 8 \| 9 |
| 3. | Country area | ☐ | Terrain ruggedness Index | ☐ | 1 \| 2 \| 3 \| 4 \| 5 \| 6 \| 7 \| 8 \| 9 |

For the Key-Indicator Earth Systems, which do you see important over the other (mark the one with a ✓) and what level of importance does the preferred one hold over the other in One Health according to you?

|  | S.no. | Indicator-1 |  | Indicator-2 |  | Level of Importance |
| --- | --- | --- | --- | --- | --- | --- |
|  | 1. | Forest | ☐ | Water | ☐ | 1 \| 2 \| 3 \| 4 \| 5 \| 6 \| 7 \| 8 \| 9 |
|  | 2. | Forest | ☐ | Air | ☐ | 1 \| 2 \| 3 \| 4 \| 5 \| 6 \| 7 \| 8 \| 9 |
|  | 3. | Forest | ☐ | Natural Disasters | ☐ | 1 \| 2 \| 3 \| 4 \| 5 \| 6 \| 7 \| 8 \| 9 |
|  | 4. | Water | ☐ | Air | ☐ | 1 \| 2 \| 3 \| 4 \| 5 \| 6 \| 7 \| 8 \| 9 |
| 5. | | Water | ☐ | Natural Disasters | ☐ | 1 \| 2 \| 3 \| 4 \| 5 \| 6 \| 7 \| 8 \| 9 |
| 6. | | Natural Disasters | ☐ | Air | ☐ | 1 \| 2 \| 3 \| 4 \| 5 \| 6 \| 7 \| 8 \| 9 |
| 7. | | Land | ☐ | Forest | ☐ | 1 \| 2 \| 3 \| 4 \| 5 \| 6 \| 7 \| 8 \| 9 |
|  | 8. | Land | ☐ | Water | ☐ | 1 \| 2 \| 3 \| 4 \| 5 \| 6 \| 7 \| 8 \| 9 |
|  | 9. | Land | ☐ | Air | ☐ | 1 \| 2 \| 3 \| 4 \| 5 \| 6 \| 7 \| 8 \| 9 |
|  | 10. | Land | ☐ | Natural Disasters | ☐ | 1 \| 2 \| 3 \| 4 \| 5 \| 6 \| 7 \| 8 \| 9 |

For the category Extrinsic drivers indices, which do you see important over the other (mark the one with a ✓) and what level of importance does the preferred one hold over the other in One Health according to you?

|  | S.no. | Key- Indicator-1 |  | Key- Indicator-2 |  | Level of Importance |
| --- | --- | --- | --- | --- | --- | --- |
|  | 1. | Earth System | ☐ | Institutional System | ☐ | 1 \| 2 \| 3 \| 4 \| 5 \| 6 \| 7 \| 8 \| 9 |
|  | 2. | Earth System | ☐ | Economic System | ☐ | 1 \| 2 \| 3 \| 4 \| 5 \| 6 \| 7 \| 8 \| 9 |
|  | 3. | Earth System | ☐ | Natural Disasters | ☐ | 1 \| 2 \| 3 \| 4 \| 5 \| 6 \| 7 \| 8 \| 9 |
|  | 4. | Earth System | ☐ | Sociological System | ☐ | 1 \| 2 \| 3 \| 4 \| 5 \| 6 \| 7 \| 8 \| 9 |
| 5. | | Earth System | ☐ | Technological System | ☐ | 1 \| 2 \| 3 \| 4 \| 5 \| 6 \| 7 \| 8 \| 9 |
| 6. | | Institutional System | ☐ | Economic System | ☐ | 1 \| 2 \| 3 \| 4 \| 5 \| 6 \| 7 \| 8 \| 9 |
| 7. | | Institutional System | ☐ | Natural Disasters | ☐ | 1 \| 2 \| 3 \| 4 \| 5 \| 6 \| 7 \| 8 \| 9 |
|  | 8. | Institutional System | ☐ | Sociological System | ☐ | 1 \| 2 \| 3 \| 4 \| 5 \| 6 \| 7 \| 8 \| 9 |
|  | 9. | Institutional System | ☐ | Technological System | ☐ | 1 \| 2 \| 3 \| 4 \| 5 \| 6 \| 7 \| 8 \| 9 |
|  | 10. | Economic System | ☐ | Natural Disasters | ☐ | 1 \| 2 \| 3 \| 4 \| 5 \| 6 \| 7 \| 8 \| 9 |
|  | 11. | Economic System | ☐ | Sociological System | ☐ | 1 \| 2 \| 3 \| 4 \| 5 \| 6 \| 7 \| 8 \| 9 |
|  | 12. | Economic System | ☐ | Technological System | ☐ | 1 \| 2 \| 3 \| 4 \| 5 \| 6 \| 7 \| 8 \| 9 |
|  | 13. | Natural Disasters | ☐ | Sociological System | ☐ | 1 \| 2 \| 3 \| 4 \| 5 \| 6 \| 7 \| 8 \| 9 |
|  | 14. | Natural Disasters | ☐ | Technological System | ☐ | 1 \| 2 \| 3 \| 4 \| 5 \| 6 \| 7 \| 8 \| 9 |
|  | 15. | Sociological System | ☐ | Technological System | ☐ | 1 \| 2 \| 3 \| 4 \| 5 \| 6 \| 7 \| 8 \| 9 |

For reference we have given the key indicators and indicators in the particular categories for assistance. For further information about the sub indicators please refer to the indicator names sheet.

| S.no. | **Categories** | Key Indicator | Component Indicators |
| --- | --- | --- | --- |
| 1. | **Extrinsic Driver index** | Earth system | Land, Forest, Water, Air, Natural disasters |
|  |  | Institutional system | Justice, Governance |
|  |  | Economic system | Finance, Work, Housing |
|  |  | Sociological system | Demography, Education, Inequalities |
|  |  | Technological system | Transport, Technology adoption, Consumption, and production |
| 2. | **Intrinsic driver index** | Human Health | Reproductive, maternal, new-born and child health, Infectious diseases, non-communicable diseases and mental health, Injuries and violence, Universal health coverage and health systems, Health risk |
|  |  | Animal health and ecosystem diversity | Animal epidemic disease, Animal welfare, Animal nutritional status, Animal biodiversity |
|  |  | Environment Health | Air quality and climate change, Land resources, Sanitation and water resources, Hazardous chemicals, Environmental biodiversity |
| 3. | **Core Drivers Index** | Governance | Participation, Rule of law, Transparency, Responsiveness, Consensus oriented, Equity and inclusiveness, Effectiveness and efficiency, Political support, |
|  |  | Zoonotic diseases | Source of infection, Route of transmission, Targeted population, Capacity building, Outcomes (case-studies) |
|  |  | Food security | Food demand and supply, Food safety, Nutrition, Natural and social circumstances, Government support and response |
|  |  | Antimicrobial resistance | Amr surveillance system, AMR laboratory network and coordination capacity, antimicrobial control and optimization, improve awareness and understanding, antimicrobial resistance rate for important antibiotics |
|  |  | Climate change | Government response, Climate change risks, Health outcome |

For the One Health Index, among the three categories in which One Health Index has been divided, which do you see important over the other (mark the one with a ✓) and what level of importance does the preferred one hold over the other in One Health according to you?

| S.no. | Key- Indicator-1 |  | Key- Indicator-2 |  | Level of Importance |
| --- | --- | --- | --- | --- | --- |
| 1. | Extrinsic Driver index | ☐ | Intrinsic driver index | ☐ | 1 \| 2 \| 3 \| 4 \| 5 \| 6 \| 7 \| 8 \| 9 |
| 2. | Extrinsic Driver index | ☐ | Core Drivers Index | ☐ | 1 \| 2 \| 3 \| 4 \| 5 \| 6 \| 7 \| 8 \| 9 |
| 3. | Intrinsic driver index | ☐ | Core Drivers Index | ☐ | 1 \| 2 \| 3 \| 4 \| 5 \| 6 \| 7 \| 8 \| 9 |

Please have a look at the list of all Categories, Key Indicators, Indicators and Sub-Indicators and give us the suggestions of which variable you feel is important and needs to be added to our lists which will be important to measure One Health apart from the available ones.

| **Category** | **Key Indicator** | **Indicator** | **Sub-indicator** |
| --- | --- | --- | --- |
| External drivers index (EDI) | Earth system | Land | Country area |
|  |  |  | Cultivated area |
|  |  |  | Arable land area |
|  |  |  | Terrain ruggedness index |
|  |  | Forest | Forest area |
|  |  |  | Forest transition phase |
|  |  |  | Trees cover (loss) |
|  |  |  | Permanent deforestation |
|  |  | Water | Renewable water resources |
|  |  |  | Water dependency ratio |
|  |  |  | Water stress |
|  |  | Air | Co₂ emissions |
|  |  |  | Air pollution index |
|  |  | Natural disasters | Disasters death rate |
|  |  |  | Disaster economic loss |
|  |  |  | Disasters affected population |
|  | Institutional system | Justice | Unsentenced detainees |
|  |  |  | Property rights |
|  |  |  | Corruption perception index |
|  |  |  | Press freedom index |
|  |  |  | Affordability of justice |
|  |  | Governance | Voice and accountability |
|  |  |  | Government spending |
|  |  |  | Public social expenditure |
|  |  |  | Public education expenditure |
|  |  |  | Public health expenditure |
|  |  |  | Political stability |
|  |  |  | Government effectiveness |
|  |  |  | Regulatory quality |
|  |  |  | Rule of law |
|  |  |  | Control of corruption |
|  | Economic system | Finance | GDP |
|  |  |  | GDP deflator |
|  |  |  | Revenue excluding grants |
|  |  |  | grants and other revenues |
|  |  |  | Adjusted GDP growth |
|  |  | Work | Labor force participation |
|  |  |  | Unemployment |
|  |  |  | Annual working hours |
|  |  |  | Youth condition |
|  |  | Housing | Own outright |
|  |  |  | Rent at reduced/subsidized price |
|  | Sociological system | Demography | Natural population growth |
|  |  |  | Life expectancy |
|  |  |  | Child and infant mortality |
|  |  |  | Total fertility rate |
|  |  |  | Urbanization |
|  |  | Education | Education enrollment |
|  |  |  | Literacy |
|  |  |  | PISA score |
|  |  |  | Science performance |
|  |  |  | Higher education |
|  |  |  | Expenditure on research |
|  |  |  | Female graduates |
|  |  |  | Researchers’ population |
|  |  | Inequalities | Gini coefficient |
|  |  |  | Palma ratio |
|  |  |  | Human development index |
|  |  |  | Poverty rate |
|  |  |  | Gender inequality index |
|  | Technological system | Transport | Railway travel |
|  |  |  | Air travel |
|  |  | Technology adoption | Internet population |
|  |  |  | Motor vehicle ownership |
|  |  |  | Mobile cellular subscriptions |
|  |  |  | Logistics performance index |
|  |  |  | Access to electricity |
|  |  |  | Share of renewable energy |
|  |  | Consumption and production | Energy consumption |
|  |  |  | Electricity consumption |
|  |  |  | Solid waste |
|  |  |  | Electronic waste |
|  |  |  | So₂ emissions |
|  |  |  | Nitrogen emissions |
|  |  |  | Non-recycled waste (plastic ones only) |
| Intrinsic drivers index (IDI) | Human health | Reproductive, maternal, new-born and child health | Maternal health |
|  |  |  | Neonatal health |
|  |  |  | Child health |
|  |  |  | Adolescent fertility |
|  |  | Infectious diseases | Tuberculosis |
|  |  |  | HIV |
|  |  |  | Malaria |
|  |  |  | Neglected tropical diseases |
|  |  |  | COVID-19 |
|  |  | Non-communicable diseases and mental health | Cardiovascular disease |
|  |  |  | Neoplasms |
|  |  |  | Diabetes mellitus |
|  |  |  | Chronic respiratory disease |
|  |  |  | Suicide |
|  |  | Injuries and violence | Road traffic |
|  |  |  | Unintentional poisoning |
|  |  |  | Homicide |
|  |  | Universal health coverage and health systems | Health coverage |
|  |  |  | Research and development Expenditure on health issues |
|  |  |  | Domestic health expenditure |
|  |  |  | Infants’ vaccination |
|  |  | Health risk | Unsafe or unimproved water, sanitation and hygiene |
|  |  |  | Household air pollution |
|  |  |  | Occupational risks |
|  | Animal health and ecosystem diversity | Animal epidemic disease | Diseases of domestic animal |
|  |  |  | Diseases of wild animal |
|  |  | Animal welfare | Overexploited or collapsed stocks Fish |
|  |  |  | Trawling or dredging fish |
|  |  |  | Discarded fish |
|  |  | Animal nutritional status | Chicken meat production efficiency |
|  |  |  | Pig meat production efficiency |
|  |  |  | Cattle production efficiency |
|  |  |  | Cattle milk production efficiency |
|  |  | Animal biodiversity | Endemic mammal species |
|  |  |  | Endemic bird species |
|  |  |  | Endemic amphibian species |
|  |  |  | Endemic reef-forming coral species |
|  |  |  | Endemic freshwater crab species |
|  |  |  | Endemic shark and ray species |
|  | Environmental health | Air quality and climate change | Ambient particulate matter pollution |
|  |  |  | Household solid fuels |
|  |  |  | Ambient ozone pollution |
|  |  |  | Climate risk |
|  |  |  | Greenhouse gas |
|  |  | Land resources | Area at risk elevation |
|  |  |  | Tree cover loss |
|  |  |  | Grassland loss |
|  |  |  | Wetland loss |
|  |  |  | Mineral depletion |
|  |  | Sanitation and water resources | Freshwater |
|  |  |  | Clean drinking water |
|  |  |  | Renewable internal freshwater resources |
|  |  | Hazardous chemicals | Fertilizer consumption |
|  |  |  | Controlled solid waste |
|  |  |  | So_2_ growth |
|  |  |  | Nox growth |
|  |  |  | Wastewater treatment |
|  |  |  | Electronic waste |
|  |  |  | Non-recycled municipal solid waste |
|  |  | Environmental biodiversity | Protected areas representativeness |
|  |  |  | Species habitat |
|  |  |  | Biodiversity habitat |
| Core drivers index (CDI) | Governance | Participation | Global connectivity |
|  |  |  | Risk communication |
|  |  |  | One Health association |
|  |  |  | One Health forums |
|  |  | Rule of law | General rule of law |
|  |  |  | One Health specialized law & regulation |
|  |  | Transparency | Transparency |
|  |  | Responsiveness | Emergency response operation |
|  |  |  | Exercising response plans |
|  |  | Consensus oriented | Consensus oriented |
|  |  |  | One Health education |
|  |  | Equity and inclusiveness | Zoonotic disease governance |
|  |  |  | Protected areas representativeness |
|  |  |  | Sustainable nitrogen management |
|  |  | Effectiveness and efficiency | Government effectiveness |
|  |  | Political support | One Health official department |
|  |  |  | Control of corruption |
|  |  |  | Regulatory quality |
|  |  |  | Government spending |
|  | Zoonotic diseases | Source of infection | Strategy and regulation |
|  |  |  | Surveillance and response |
|  |  |  | Sanitation |
|  |  | Route of transmission | Detection |
|  |  |  | Vector and reservoir interventions |
|  |  | Targeted population | Vaccination regulation |
|  |  |  | Population coverage and cost of interventions |
|  |  |  | Inhabitants below 5 meters above sea level |
|  |  | Capacity building | Zoonosis health promotion |
|  |  |  | Natural protected areas |
|  |  | Outcomes (case-studies) | COVID-19 confirmed cases |
|  |  |  | Human DALYs of echinococcosis |
|  |  |  | Human DALYs of leishmaniasis |
|  |  |  | Human DALYs of rabies |
|  |  |  | Human DALYs of tuberculosis |
|  | Food security | Food demand and supply | Food demand score (SECURITY) |
|  |  |  | Food loss and waste |
|  |  |  | Infrastructures score |
|  |  |  | Food import score |
|  |  |  | Food production score |
|  |  | Food safety | Food safety governance |
|  |  |  | Food quality control |
|  |  |  | Food safety score |
|  |  |  | Foodborne illness burden |
|  |  | Nutrition | Food balance |
|  |  |  | Nutrition promoting capacity |
|  |  |  | Nutrition score |
|  |  | Natural and social circumstances | Famine warning |
|  |  |  | Natural sources sustainability in Land and Water |
|  |  |  | Economic performance index |
|  |  |  | Agriculture value added per worker |
|  |  |  | Food price indicators |
|  |  | Government support and response | Investment and financial support Score |
|  |  |  | Training and AI agriculture performance score |
|  | Antimicrobial resistance | Amr surveillance system | Antimicrobial consumption both in human and animals |
|  |  |  | Antimicrobial resistance status in human, animal and food |
|  |  |  | Environmental surveillance system |
|  |  | AMR laboratory network and coordination capacity | National AMR capacity |
|  |  |  | Technical promotion scores in AMR |
|  |  |  | National action plan formulations |
|  |  | Antimicrobial control and optimization | National law for antibiotic use |
|  |  |  | Optimization of antimicrobial use |
|  |  |  | Interruption capacity of antimicrobial resistance transmission |
|  |  | Improve awareness and understanding | Raising awareness and understanding |
|  |  |  | Professional training activities in multi-sectors |
|  |  | Antimicrobial resistance rate for important antibiotics | Carbapenems-resistance for multi-species, e.g., *Klebsiella pneumoniae, Acinetobacter baumannii, Escherichia coli, Pseudomonas aeruginosa* |
|  |  |  | Vancomycin-resistance for *Enterococcus faecium*, and *Enterococcus faecalis* |
|  |  |  | Third-generation Β-lactams-resistance for multi-species, e.g., *Staphylococcus aureus, Klebsiella pneumoniae, Escherichia coli, Pseudomonas aeruginosa* |
|  |  |  | Macrolides-resistance for Streptococcus pneumoniae |
|  |  |  | Aminoglycosides-resistance for *Klebsiella pneumoniae and Acinetobacter baumannii* |
|  |  |  | Quinolone-resistance for *Klebsiella pneumoniae, Escherichia coli, Acinetobacter baumannii* |
|  | Climate change | Government response | Climate policy |
|  |  |  | Climate knowledge system |
|  |  |  | Climate intervention strategy |
|  |  | Climate change risks | Air condition score |
|  |  |  | Extreme weather indicators, e.g. wildfires, droughts, floods, extreme temperatures |
|  |  |  | Energy use indicators, e.g. oil, natural gas, electricity, coal |
|  |  |  | Greenhouse gas emissions score |
|  |  | Health outcome | Directly health outcome indicators |
|  |  |  | Indirectly health outcome indicators |
